# Genetic prediction of colorectal cancer risk in six major ancestries provides insights to streamline practice screening guidelines

**DOI:** 10.64898/2026.08.10.26360074

**Authors:** Abishna Parasuraman, Amanda Wei-Yin Lim, Rawaa Eltayib, Nirmala Pandeya, Catherine M Olsen, Graham Radford-Smith, David C. Whiteman, Stuart MacGregor, Mathias Seviiri

**Author notes:** Corresponding author: Mathias Seviiri, Statistical Genetics, Department of Population Health, QIMR Berghofer, Brisbane, Australia., 300 Herston Road, Herston, Brisbane, Queensland 4006, Australia.

## Abstract

**Background and objective:** Colorectal cancer (CRC) is the third leading cause of cancer deaths worldwide. Early identification of high-risk individuals allows targeted prevention and early detection.

**Design:** We constructed a polygenic risk score (PRS) for CRC risk using data from 1,448,354 individuals (103,401 cases). We evaluated its performance for identifying high-risk individuals in 6 major ancestries.

**Results:** The PRS was strongly associated with CRC risk in Europeans (OR per SD =2.13, 95%CI=1.98-2.28), Africans (OR=1.35, 95%CI=1.11-1.64), Hispanics (OR=1.97, 95%CI =1.48-2.61), East Asians (OR=1.98, 95%CI=1.35-2.91), South Asians (OR=1.85, 95%CI=1.44 - 2.37), and Middle Easterners (OR=3.10, 95%CI=1.38-6.95). Europeans in the top 10% genetic risk had 14-fold and 5-fold higher CRC risks compared to the bottom 10% (OR=13.50, 95%CI=8.67-21.00), and average (20-70%) risk groups (OR=4.64, 95%CI=3.89-5.52), respectively. The CRC risk in the top 10% individuals was equivalent to having three affected first degree relatives with CRC diagnosed at any age. Genetically high-risk individuals developed CRC up to 15 years earlier than the average. The PRS was strongly associated with early onset CRC risk e.g. in AFR (OR=3.22, 95%CI=1.79-5.81), and improved its prediction e.g. by 9% beyond clinical predictors in EUR.

**Conclusion:** A comprehensive genetic prediction of CRC risk provides insights that could streamline screening and prevention guidelines.

**What is already known on this topic:**

- Current screening and prevention programs for CRC rely on clinical assessments of future risk. We investigated whether a genetic test (PRS) can identify high-risk individuals to be prioritised for screening.

**What this study adds:**

- The PRS identified the top 10% of individuals with 5-fold higher CRC risk and developed CRC 15 years earlier than the average, and improved the prediction of early-onset CRC by 9%.
- Individuals in the top 10% PRS (high genetic risk) with no family history of CRC had the same risk as those with at least three first degree relatives with CRC diagnosed at any age.

**How this study might affect research, practice or policy:**

- Our findings provide comprehensive evidence to support potential integration of polygenic screening into clinical practice guidelines for CRC screening, early identification and prevention across different populations or ancestries

## INTRODUCTION

Colorectal cancer (CRC) is the third leading cause of cancer death worldwide, with incidence and mortality doubling over two decades to ∼2.17 million cases and 1.09 million deaths annually respectively [1]. Rates are rising in individuals <50 years [2], yet screening programs mainly target those ≥50 or younger individuals with family history or symptoms. In the United States of America (USA) and Australia, ages 45–49 can only opt in [3–6]. Limited screening leads to later-stage diagnosis and poorer survival, highlighting the urgent need to identify individuals at high risk of early-onset CRC (EOCRC) or those with no family history that do not meet current clinical criteria for screening.

GWAS have identified many common CRC susceptibility variants [7], highlighting a strong genetic component and motivating the need for polygenic screening using polygenic risk score (PRS). CRC PRSs can identify high-risk individuals [8,9] and have been proposed for clinical use [10], but most studies are Euro-centric and perform sub-optimally in non-European populations [11], limiting translation. PRS evaluation in individuals <50 years is also limited. Yet, genetic screening at any age could enable early risk identification, personalized prevention, and re-stratification of clinically high-risk patients (for example with family history) for prioritization for diagnostic assessment or surveillance amidst the long waiting list.

In this study, we leveraged large scale clinical and genetic data from a total of 1.94 million individuals including 115,849 CRC cases to construct a CRC PRS using new and powerful PRS methods. We evaluated its ability to identify individuals with a high genetic risk for both “any age” CRC and EOCRC in both European and non-European populations, and examined its ability to stratify risk beyond traditional risk factors. We provide suggestions in relation to the current screening guidelines and on how genetic screening could refine the current CRC screening and prevention strategies.

## METHODS AND MATERIALS

### Study populations and settings

#### Discovery cohorts for developing the polygenic risk score

We collated genetic and clinical data from large biobanks in Australia, the USA, Canada, Europe, and Asia to conduct the discovery GWAS for PRS development **(Figure 1; Supplementary Table 1**). Major sources included the National Institutes of Health’s All of Us Research Program (AoU) [12,13], the UK Biobank [14,15], FinnGen [16], the Million Veteran Program (MVP) [17,18], and the Canadian Longitudinal Study on Aging (CLSA) [19,20], and published GWAS from the Genetics & Epidemiology of Colorectal Cancer Consortium (GECCO) and The Colorectal Transdisciplinary Study (CORECT) consortia [7]. Non-overlapping samples were used for validation. Multi-ancestry modeling used additional GWAS from MVP (AFR, AMR) and East Asian cohorts; Biobank Japan (BBJ) [21], National Biobank of Korea (NBK) [22], and The China Kadoorie Biobank (CKB) [23].

**Figure 1:**
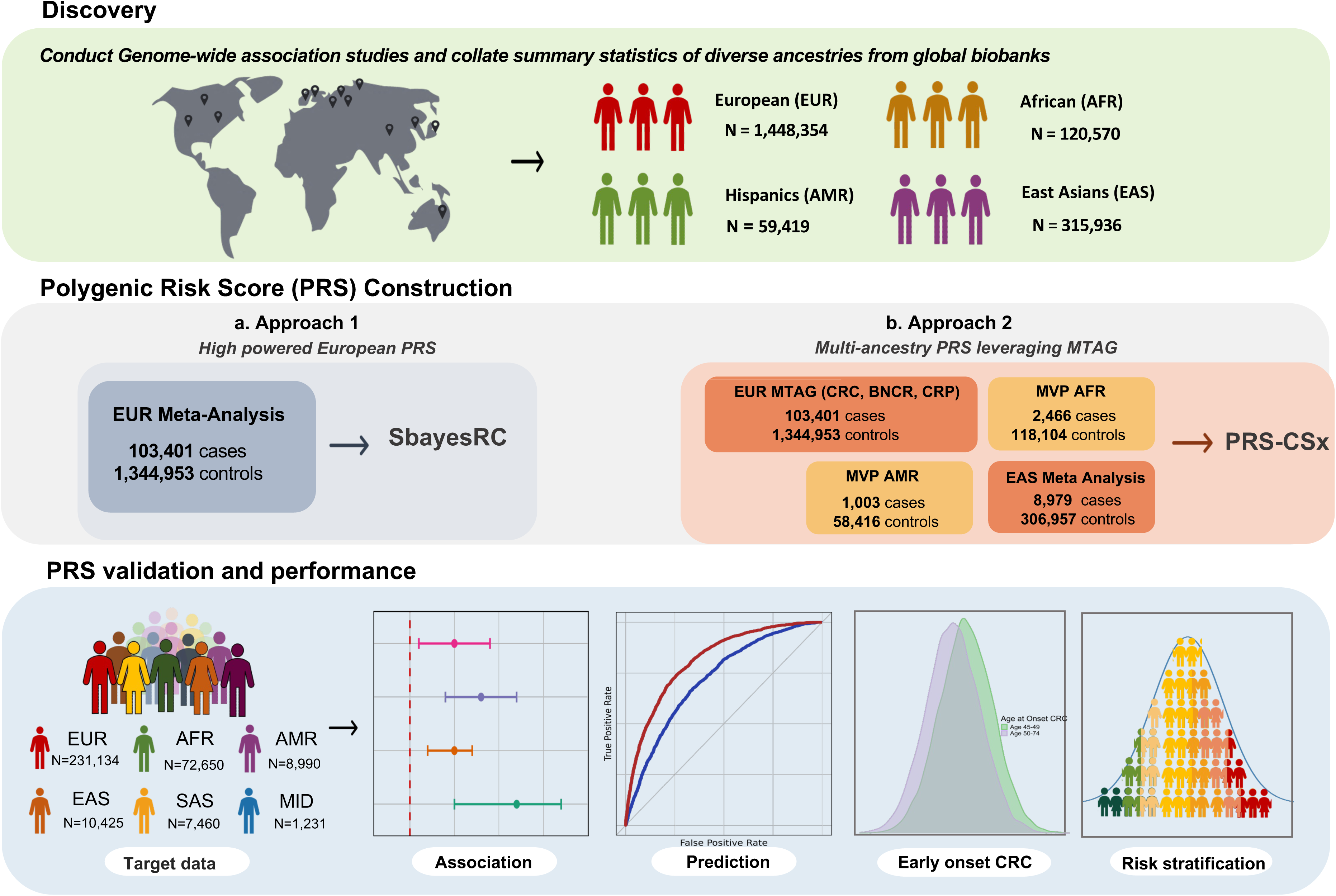
Top panel: The discovery data sets included large scale genome-wide association studies of colorectal cancer (CRC) from global biobanks on four major ancestries. Middle panel: PRS constructed with two approaches; Approach 1 leveraging European meta-analysis with functional annotations using SBayesRC, and Approach 2 leveraging European multi trait analysis GWAS (MTAG) of CRC, colorectal polyps, and benign neoplasm of colon and rectum, with a multi-ancestry PRS method, PRS-CSx. Bottom panel: Analysis performed in independent target data, of EUR, AFR, AMR, EAS, South Asian (SAS), and Middle Eastern (MID) individuals: association testing, assessing model prediction, analyzing early onset CRC, and stratifying risk across genetic strata including comparisons with family history.

#### Cohorts for evaluating the performance of the polygenic risk score

We evaluated the PRS across six ancestries in UK Biobank [14,15] (EUR, AFR, AMR, EAS, SAS), QSkin [24] (EUR), All of Us [12,13] (EUR, AFR, AMR, EAS, MID), and CARTaGENE [25,26] (EUR). Cohort details are provided in the **Supplementary Methods**. All contributing cohorts received appropriate ethical approvals, and all participants provided informed consent to participate in the study (**Supplementary Methods).** The human research ethics committee at QIMR Berghofer Medical Research Institute provided ethical oversight to this study (P3501).

### Statistical analyses

#### PRS construction approach 1: SBayesRC

We conducted a large trans-biobank GWAS meta-analysis in EUR (N = 1,448,354; 103,401 cases) (**Figure 1, Supplementary Methods,** and **Supplementary Table 1)** and constructed a “main PRS” using SBayesRC [27], which leverages ∼7 million SNPs, linkage disequilibrium structure, and functional annotations. Two additional PRSs were built: one trained in 90% of UKB EUR and tested in the 10% holdout, and one excluding AoU and tested in AoU. Raw PRS scores were generated with PLINK [28] and normalized within each validation cohort (QSkin, UKB, CARTaGENE, AoU).

#### PRS construction approach 2: PRS-CSx

We collated case-control GWAS data from 1,944, 279 individuals (115,849 CRC cases) that included EUR (N=1,448,354; 103,401 cases), EAS (N=315,936; 8979 cases), AFR (N=120,570; 2,466 cases) and AMR (N=59,419; 1,003 cases) (**Figure 1, Supplementary Methods,** and **Supplementary Table 1)**. Next, we constructed a trans-ancestry PRS using PRS-CSx [29], which jointly models ancestry-specific LD and effects sizes to improve cross-population prediction. Based on PRS weights from PRS-CSx, PLINK 1.9/2.000 [28] was used to generate the raw PRS scores for each individual in the validation datasets of non-European ancestry in the UKB, and AoU.

#### Assessing the association between the PRS and CRC risk in European and non-Europeans

First, we assessed the association between the PRS and the CRC risk by fitting a logistic regression to estimate the odds ratio per standard deviation increase in the PRS (OR per SD) and 95% confidence intervals (95% CI). We adjusted for age, sex, and 10 PCs. We assessed the associations in UKB (EUR, AFR, AMR, EAS and SAS), AoU (EUR, AFR, EAS, AMR, and MID), QSkin (EUR) from Australia and CARTaGENE (EUR) from Canada.

### Evaluating risk stratification potential of the PRS in European and non-Europeans

We evaluated PRS stratification of CRC risk by estimating odds ratios (ORs) and 95% CIs across genetic risk strata (deciles, quintiles) using multiple reference groups (bottom decile and 20–70th centile) in EUR and non-EUR populations, with and without family history. Using UKB, we also estimated the average age at CRC onset by genetic risk relative to the average genetic risk (50th centile) in EUR, AFR, EAS, and AMR. All models were adjusted for age, sex, and 10 PCs.

#### Evaluation of the predictive ability of the PRS over the clinical and lifestyle factors in Europeans and non-Europeans

Most CRC screening guidelines rely primarily on age and family history [3–6,30]. We evaluated the incremental predictive value of the PRS beyond conventional factors by adding it to models including age, sex, and 10 PCs. Using the pROC package, we computed and compared AUCs for clinical and genetics-based models across UKB, AoU, QSkin, and CARTaGENE in multiple ancestries. We further assessed AUC improvement when adding the PRS to models including first-degree family history, smoking, and inflammatory bowel disease.

#### Evaluating the potential of the PRS to reclassify individuals with affected first degree relatives for CRC risk in Europeans

CRC guidelines classify individuals with affected first-degree relatives (FDRs) as high risk and prioritize them for screening [31]. We evaluated whether the PRS could further stratify these individuals to refine screening prioritization. In UK Biobank (EUR), we estimated and compared the lifetime absolute risk for CRC across PRS centiles for individuals with at least one or two affected FDRs, and with no family history. Second, we identified the proportion of those with no family history whose risk matched that of individuals with one or two affected FDRs at average (50th centile) genetic risk. Finally, we mapped genetic risk profiles in individuals without family history to CRC risk categories defined in the 2023 Australian screening guidelines [31].

#### Assessing the association and prediction of early-onset CRC in Europeans and non-Europeans

Current national screening guidelines for CRC in Australia and the USA allow individuals 45-49 years to opt in for screening rather than wait until they are 50 years of age [5,6]. In the AoU we assessed the association of the PRS with the risk of EOCRC in individuals i) aged 45-49: EUR (N=11,595; 55 cases), AFR (N=5,604; 16 cases) and AMR (N=6,292; 21 cases), and ii) < 50 years. Next, we compared the improvement in the prediction of CRC in 45-49 vs 50+ year olds. Lastly, we tested the risk stratification ability of the PRS for EOCRC by comparing the odds of developing CRC in different genetic risk strata (e.g. top 10%) compared to those with an average genetic risk (20-70th centile).

#### Patient and Public Involvement

There was no patient or the public involvement in the design, or conduct, or reporting, or dissemination of our study.

## RESULTS

### Descriptive characteristics of the discovery and target populations

Detailed descriptive characteristics of the participants in all the target cohorts for each ancestry are presented in **Supplementary Table 2**. For example, analysis in QSkin was restricted to participants of EUR descent, on average (standard deviation (sd)) aged 58.53 (9.79) years at the time of enrolment, with the majority (57.37) being females. This was similar to the EUR participants CARTaGENE who were 54.31 (7.86) old on average at baseline recruitment, and dominated by females (53.31%).

### The PRS was strongly associated with the risk of CRC in European, Hispanic, African, Asian and Middle Eastern populations

After adjusting for age, sex, and the first 10 PCs, one standard deviation (SD) increase in the genetic risk or PRS was strongly associated with CRC risk across populations of European (EUR) descent in the United Kingdom, United States, Australia and Canada (**Figure 2A, Supplementary Table 3, and Supplementary Table 4)** e.g. UKB OR per SD = 2.13, 95% CI = 1.98 - 2.28; P=2.95 x 10^−93^. A strong association was still observed after further adjustment for the first degree relative-family history of CRC, smoking status, and IBD diagnosis (OR per SD = 2.11, 95% CI = 1.96 - 2.27; P=9.50 x 10^−90^).

**Figure 2:**
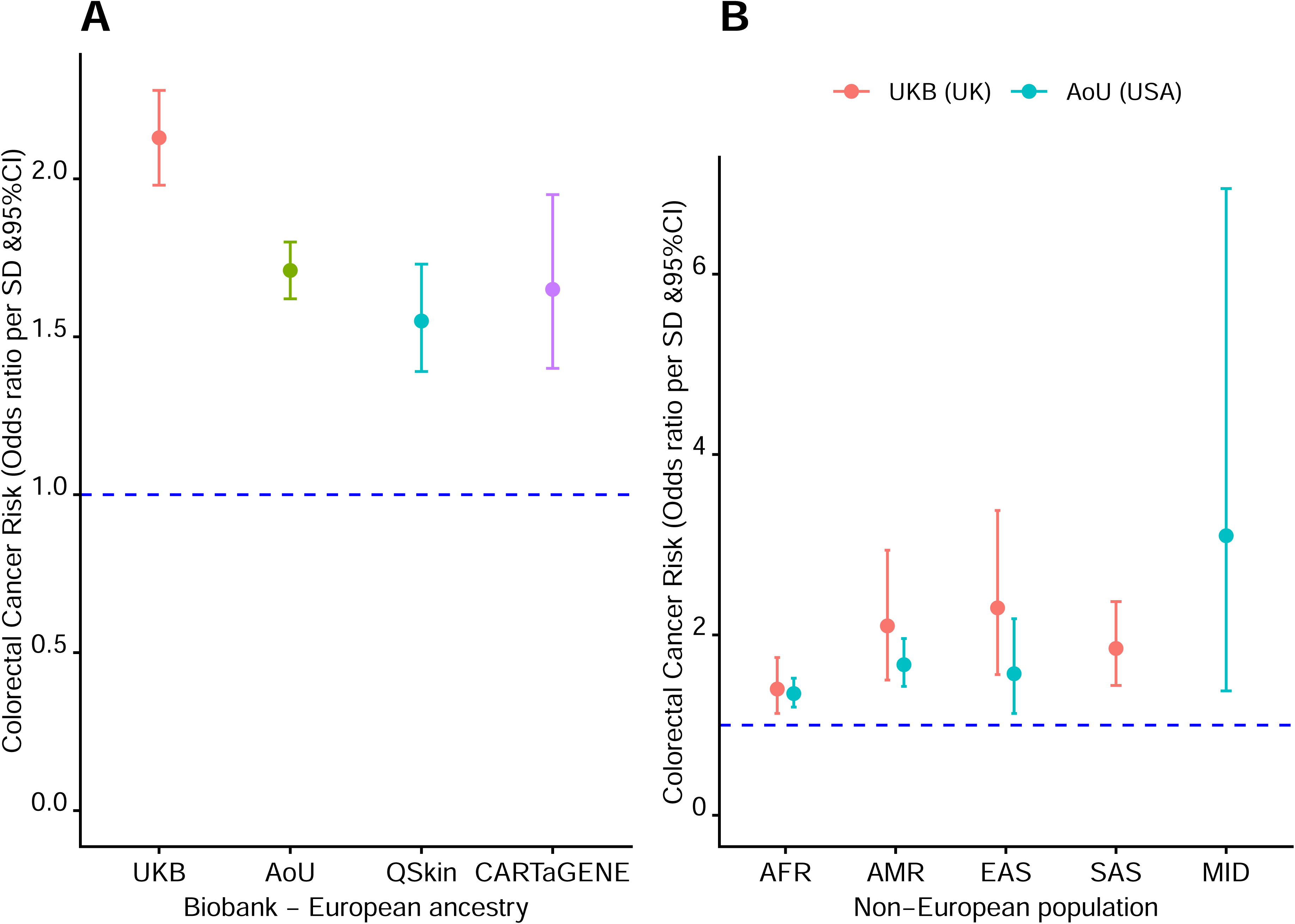
Association of the polygenic risk score and the risk of colorectal cancer in both Europeans and non-European populations. **Panel A:** Association of the polygenic risk score and the risk of colorectal cancer in European populations in the UK Biobank (United Kingdom), All of Us Research Program (the United States of America - USA), QSkin (Australia), and CARTaGENE (Canada). X-axis shows the different biobanks with participants of European ancestry. Y-axis represents the odds ratios per standard deviation (SD) increase in the genetic risk and 95% confidence intervals (CI), adjusted for age, sex, and 10 principal components. **Panel B:** Association of the polygenic risk score and the risk of colorectal cancer in non-European populations in the UK Biobank (United Kingdom), and All of Us Research Program (the United States of America - USA). X-axis shows the different ancestries - African (AFR), Hispanics (AMR), East Asian (EAS), South Asians (SAS) and Middle Easterners (MID). Y-axis represents the odds ratios per standard deviation (SD) increase in the genetic risk and 95% confidence intervals (CI), adjusted for age, sex, and 10 principal components. Red and cyan represent the UK Biobank and All of US cohorts respectively.

One SD increase in the genetic risk was strongly associated with CRC risk in populations of non-European ancestries including; Africans (UKB OR per SD =1.35, 95% CI = 1.11 - 1.64, P=2.27 x 10^−3^, and AoU OR per SD = 1.35, 95% CI = 1.20 - 1.52, P=5.50 x 10^−7^), Hispanics (UKB OR per SD =1.97, 95% CI = 1.48 - 2.61, P=2.60 x 10^−06^, and AoU OR per SD = 1.67, 95% CI = 1.43 - 1.96, P=1.53 x 10^−10^), East Asians (UKB OR per SD =1.98, 95% CI = 1.35 - 2.91, P=4.99 x 10^−4^, and AoU OR per SD = 1.57, 95% CI = 1.13 - 2.18, P=7.09 x 10^−3^), South Asians (UKB OR per SD =1.85, 95% CI = 1.44 - 2.37, P=1.26 ×10^−6^), and Middle Easterns (AoU OR per SD = 3.10, 95% CI = 1.38 - 6.95; P=5.97 x 10^−3^) (**Figure 2B**, **Supplementary Table 5, Supplementary Table 6**, and **Supplementary Table 7).** The SBayesRC and PRS-CSx models performed almost equally well (**Supplementary Table 5, Supplementary Table 6**, and **Supplementary Table 7).**

### The PRS identified individuals who might benefit from early genetic screening

The PRS identified 10% of the EUR population with 14-fold increased risk of developing CRC (OR=13.50, 95%CI=8.67-21.00, P=8.72×10^−31^) including the ultra-high risk (top 1%) individuals with 23-fold increased risk (OR=22.92, 95%CI=12.80-41.06, P=6.07×10^−26^), compared to individuals with low genetic risk (bottom 10%) in the UKB (**Figure 3 A** and **Supplementary Table 3)**. The risk was twice as high in the UKB compared to the other EUR cohorts e.g. in AOU (OR = 5.35, 95% CI = 4.19 - 6.83, P=1.62×10^−41^), CARTaGENE (OR = 4.62, 95% CI = 2.02 - 10.58, P=2.94×10^−04^) and QSkin (OR= 4.63, 95% CI = 2.58 - 8.31, P=2.84×10^−07^) (Supplementary Table 3 and Supplementary Table 4).

**Figure 3:**
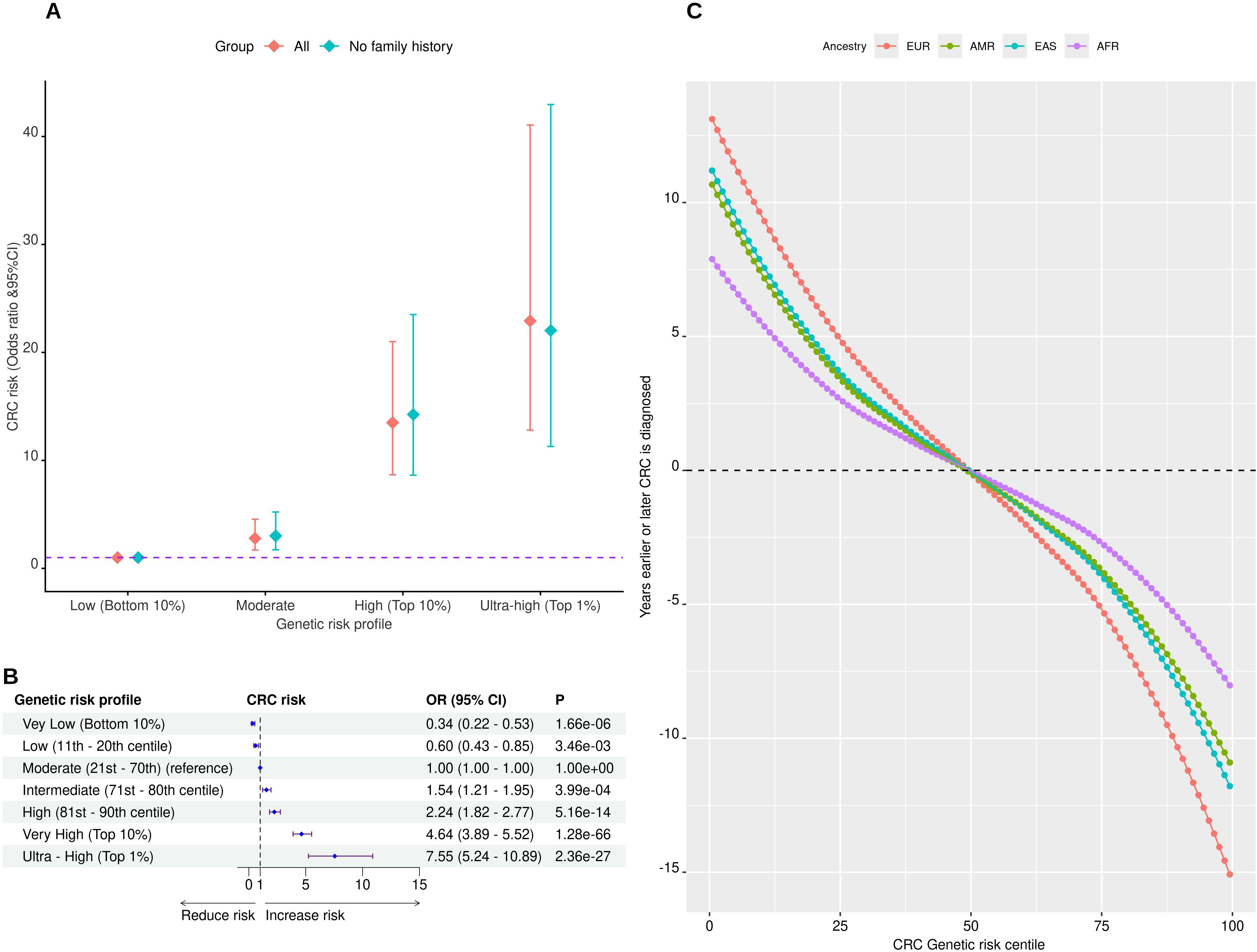
Risk stratification of the participants in the UK Biobank by the polygenic risk score (PRS). **Panel A:** Stratification of the European participants by the PRS. The x-axis represents the genetic risk profiles for colorectal cancer (CRC); low (bottom 10%), moderate (middle 10%), high (top 10%), and ultra-high (top 1%). The y-axis shows the odds ratios (OR) and 95% confidence intervals (CI) for developing colorectal cancer for individuals with moderate, high and ultra-high genetic risk compared to those with low genetic risk. Age, sex, and 10 principal components were adjusted for. The red and cyan error bars represent “all participants” and “participants without first degree relative” respectively. **Panel B:** The odds of developing colorectal cancer for individuals with very low, low, intermediate, high, very high and ultra-high genetic risk profiles, compared with those with a moderate or average risk among Europeans in UK Biobank. P refers to P-value. The vertical dashed line represents the null. **Panel C:** The relationship between the genetic risk for CRC (in centiles), and the time (in years) when participants are likely to develop CRC compared to the participants with average genetic risk (50th centile). The red, cyan, green and purple represent participants of European, East Asian, Hispanics and African ancestries in the UK Biobank.

Compared to individuals with moderate genetic risk (20th - 70th centile), EUR individuals in the top 10% had up to 5-fold increased risk of CRC (e.g. UKB OR =4.64, 95%CI = 3.89 - 5.52, P=1.28×10^−66^, and AoU OR = 3.04, 95% CI = 2.66 - 3.47, P= 5.86×10^−61^) (**Figure 3B**; **Supplementary Table 3** and **Supplementary Table 4**). Conversely, individuals with a low genetic risk (bottom 10%), had up to 66% reduced risk of CRC (e.g. UKB OR =0.34, 95%CI = 0.22 - 0.53, P=1.66×10^−06^, and AoU OR = 0.57, 95% CI = 0.45 - 0.72, P= 1.91×10^−06^) (**Figure 3B; Supplementary Table 3 and Supplementary Table 4).**

In African ancestry populations, individuals in the top 10% PRS had markedly higher CRC risk than those in the bottom 10% (e.g., UKB OR = 6.62, 95% CI = 1.46–29.91, P=1.41×10^−02^), and a 2-fold increased risk compared with average risk (20th–70th centile) (e.g. AoU OR = 2.04, 95%CI=1.48 - 2.82, P=1.60×10^−05^) (**Supplementary Table** 5). In Hispanic ancestry populations from UKB and AoU, individuals in the top 10% PRS had a 6-fold higher CRC risk than those at low genetic risk (e.g. AoU OR = 6.05, 95% CI = 3.08–11.87, P=1.64×10^−07^), and a 5-old increased risk compared with moderate risk (20th–70th centile) (OR = 5.06, 95% CI = 2.41– 10.62, P = 1.81×10□□) (**Supplementary Table 6)**.

East Asians in the top 10% PRS had up to a 4-fold increased risk of CRC, compared with moderate risk (middle 20th-70th centile) (AoU OR =3.53, 95% CI = 1.63 - 7.63, P=1.34×10^−03^) Those in the top 20% PRS had a 7-fold increased risk of CRC compared with the bottom 20% (UKB OR =7.26, 95% CI = 1.60 - 32.93, P=1.02×10^−02^) (**Supplementary Table** 7). South Asians in the UKB top 20% had a 6-fold and 3-fold increased risk of CRC compared with the bottom 20% and the 20th - 70th centile respectively (**Supplementary Table** 7).

Compared with individuals at moderate risk (50th centile), those with higher genetic risk developed CRC several years earlier—up to 15 years earlier for ultra-high–risk Europeans (top 1%) and ∼11 years earlier for ultra-high–risk Hispanics and East Asians. Conversely, low-risk individuals developed CRC later, by 10–12 years in Europeans and 7–11 years in East Asians and Hispanics in the bottom 10% (**Figure 3C**).

### The PRS improved prediction of CRC over and above the clinical risk factors

The PRS improved identification of high risk individuals over and above clinical predictors in both European and Non-European ancestry populations, e.g. by up to 7% beyond age and sex in the UKB (AUC = 0.78 vs 0.71; P < 2.2×10^−16^), QSkin (AUC = 0.71 vs 0.67; by 4%; P= 5.566×10^−05^), CARTaGENE (AUC = 0.77 vs 0.74; by 3%; P= 0.01945), and AoU (AUC = 0.79 vs 0.76; by 3%; P < 2.2×10^−16^) (**Figure 4A, and Supplementary Table 8).**

**Figure 4:**
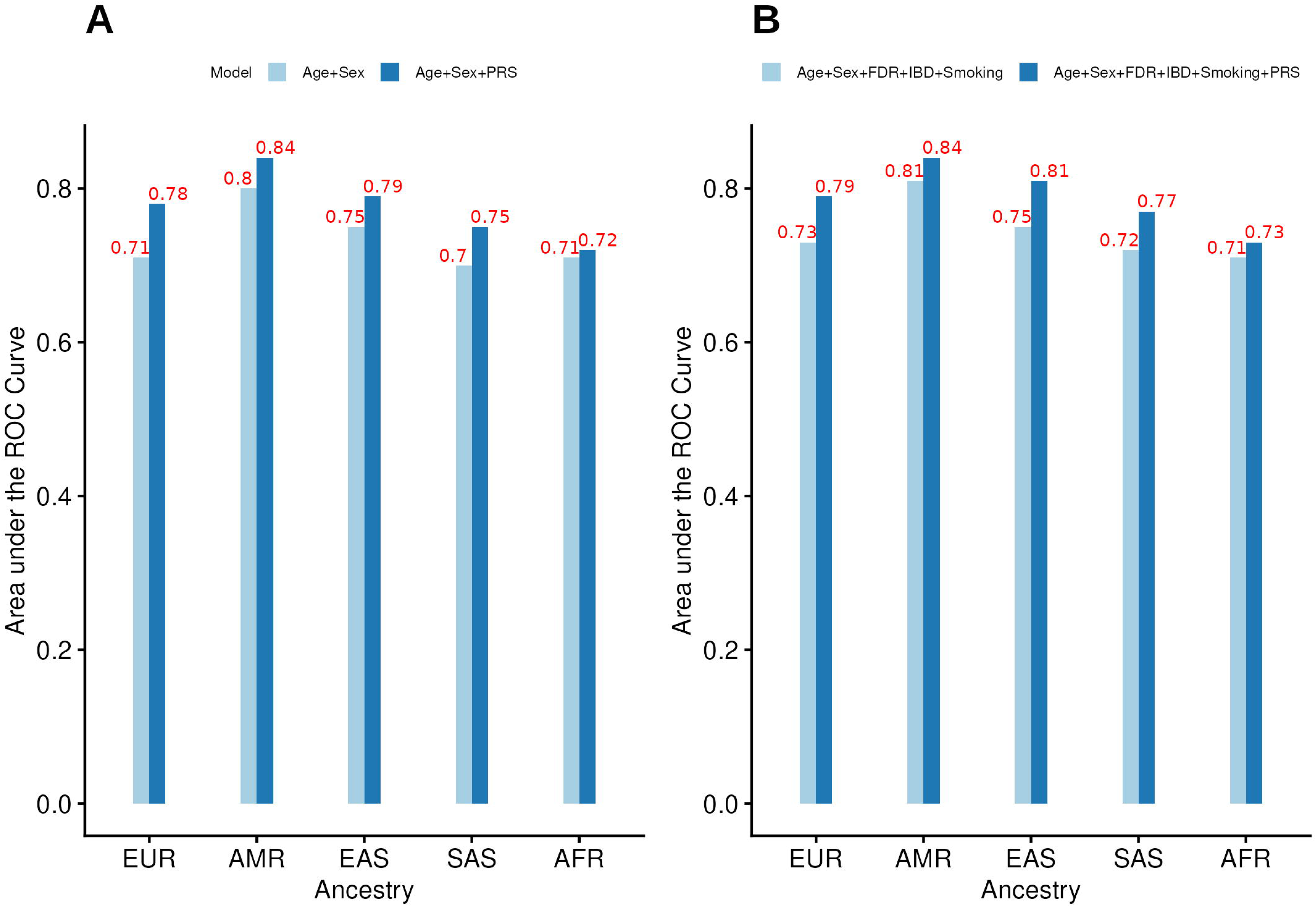
Comparison of the prediction of CRC risk for models with and without the polygenic risk score in the UK Biobank. The x-axis represents the different ancestries; EUR - Europeans, AFR - Africans, AMR - Hispanics, EAS - East Asians, and SAS - South Asians. The y-axis represents the area under the receiver operating curve on a scale of 0-1. **Panel A**: Comparison of the prediction of CRC risk for the clinical model (cyan) with age, sex, and 10 principal components, and when the PRS is added to the clinical model (blue). **Panel B**: Comparison of the prediction of CRC risk for the clinical model (cyan) with age, sex, 10 principal components, smoking, first degree relative, and inflammatory bowel disease and when the PRS is added to the clinical model (blue).

It also significantly improved the prediction of CRC risk beyond age, and sex in Hispanics by up to 4% (UKB AUC = 0.84 vs 0.80, P=0.01569, and AoU AUC = 0.79 vs 0.77, P=7.157 x 10^−4^), in East Asians by up to 4% (UKB AUC = 0.79 vs 0.75, P=0.03088, and AoU AUC = 0.80 vs 0.78, P=0.3274), and in South Asians by 5% (UKB AUC = 0.75 vs 0.70, P=0.01949) (**Panel 4A**). The improvement was not significant in Africans and Middle Easterns.

The PRS significantly improved CRC prediction beyond family history, smoking, IBD, age, and sex in UKB by 6% in EUR (AUC 0.79 vs 0.73, P < 2.2×10□¹□), 3% in AMR (0.84 vs 0.81, P = 0.017), 6% in EAS (0.85 vs 0.75, P = 0.010), and 5% in SAS (0.77 vs 0.72, P = 0.019) (**Figure 4B; Supplementary Table 8**). Improvement in AFR was 2% but not significant (0.73 vs 0.71, P = 0.176). In CARTaGENE, the PRS increased prediction by 3% beyond age, sex, and family history (0.78 vs 0.75, P = 0.021).

### The PRS reclassified individuals with first degree family history of CRC

The PRS effectively reclassified CRC risk among individuals with affected FDRs. Those in the bottom 20% PRS with an affected FDR had lifetime risk comparable to individuals without family history, whereas lifetime risk reached 25% in the top 1% with two affected FDRs (**Figure 5**). Conversely, individuals without family history in the top 10% PRS had risks comparable to or exceeding those with two affected FDRs at average genetic risk, and 20% without family history (top 20% PRS) had risks similar to or higher than those with at least one affected FDR **(Figure 5**).

**Figure 5.**
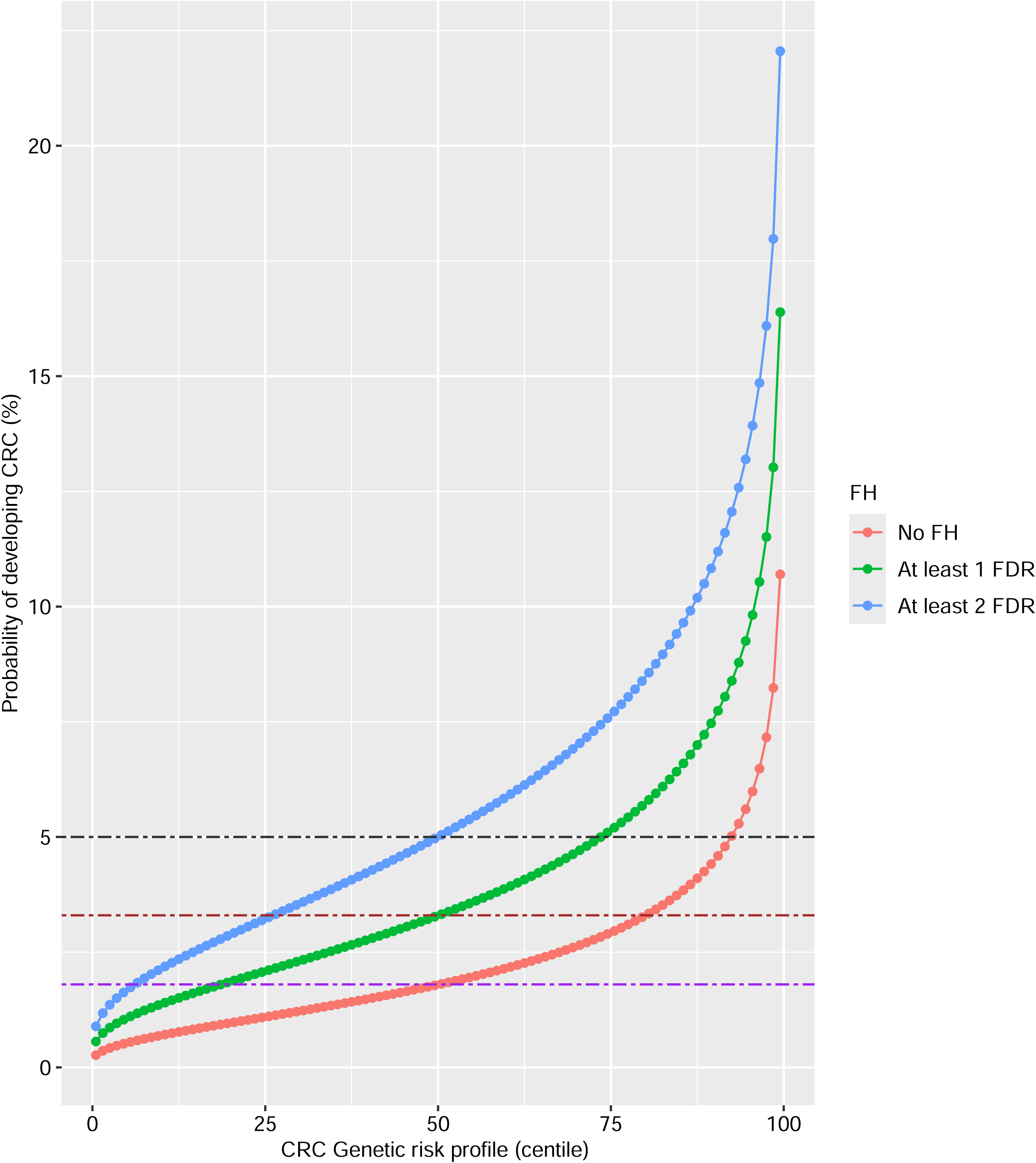
Stratification of individuals with an affected first degree relative (FDR), and without family history for colorectal cancer in the UK Biobank participants of European ancestry using the life-time absolute risk. The x-axis represents the genetic risk profile in centile calculated using the mean and standard deviation of European ancestry participants in 1000 Genomes Phase 3 data. The y-axis represents the probability of developing colorectal cancer in percentage. The red, green and blue curves represent participants without family history, with at least 1, and with at least 2 affected FDRs respectively. The horizontal purple, brown, and grey lines demonstrate absolute risks equivalent to that of people with no family history, at least 1 FDR, and at least 2 FDR but with average (50th centile).

Relative to the 2023 Australian guidelines [31], individuals without family history in the top 30% and top 10% PRS mapped to risk categories 2 and 3, respectively (**Table 1**). The PRS strongly stratified those without family history, identifying high-risk individuals otherwise classified as low clinical risk and potentially eligible for CRC screening. Those in the top 10% PRS had a 14-fold higher CRC risk than low-risk counterparts (OR = 14.25, 95% CI = 8.63– 23.51, P = 2.66×10□²□), exceeding the effect observed among individuals with affected FDRs (top 10% OR = 7.56, 95% CI = 3.31–17.30, P = 1.66×10□□) (**Figure 3A; Supplementary Table 9**).

**Table 1:**
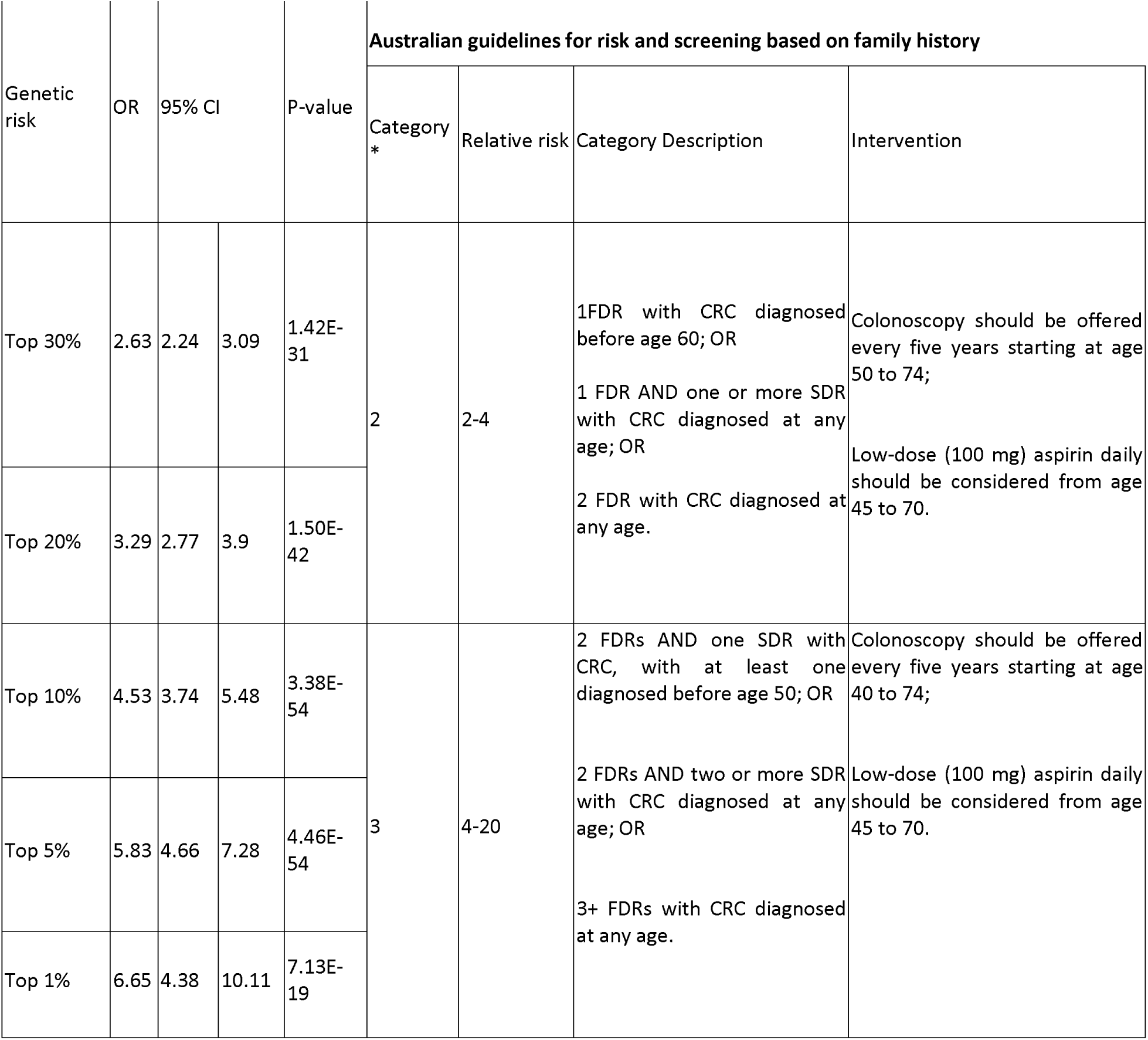
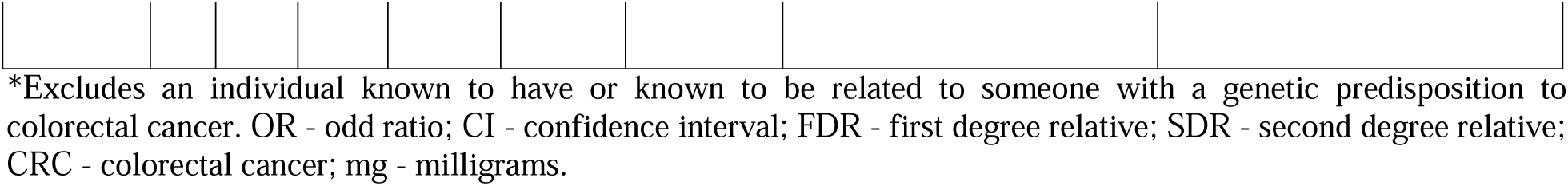
Comparison of the genetic risk profile for participants with no family history to the equivalent risk category for screening based on family history from the Australian guidelines.

### The PRS was strongly associated with and predicted early onset of CRC

Updated CRC national screening guidelines include individuals 45-49 years to opt in for testing in Australia and USA. Across EUR, AFR, and AMR in AoU, the PRS showed stronger associations with EOCRC (45–49 years) than with late-onset CRC (LOCRC, ≥50 years). In EUR, the per-SD effect was ∼30% higher for EOCRC (OR = 2.03, 95% CI = 1.55–2.66, P=2.76 x 10^−7^) than for LOCRC (OR = 1.69, 95% CI = 1.60–1.78, P = 2.25 x 10^−8^) (**Figure 6A**), with greater risk stratification for EOCRC (**Figure 6B**).

**Fig 6:**
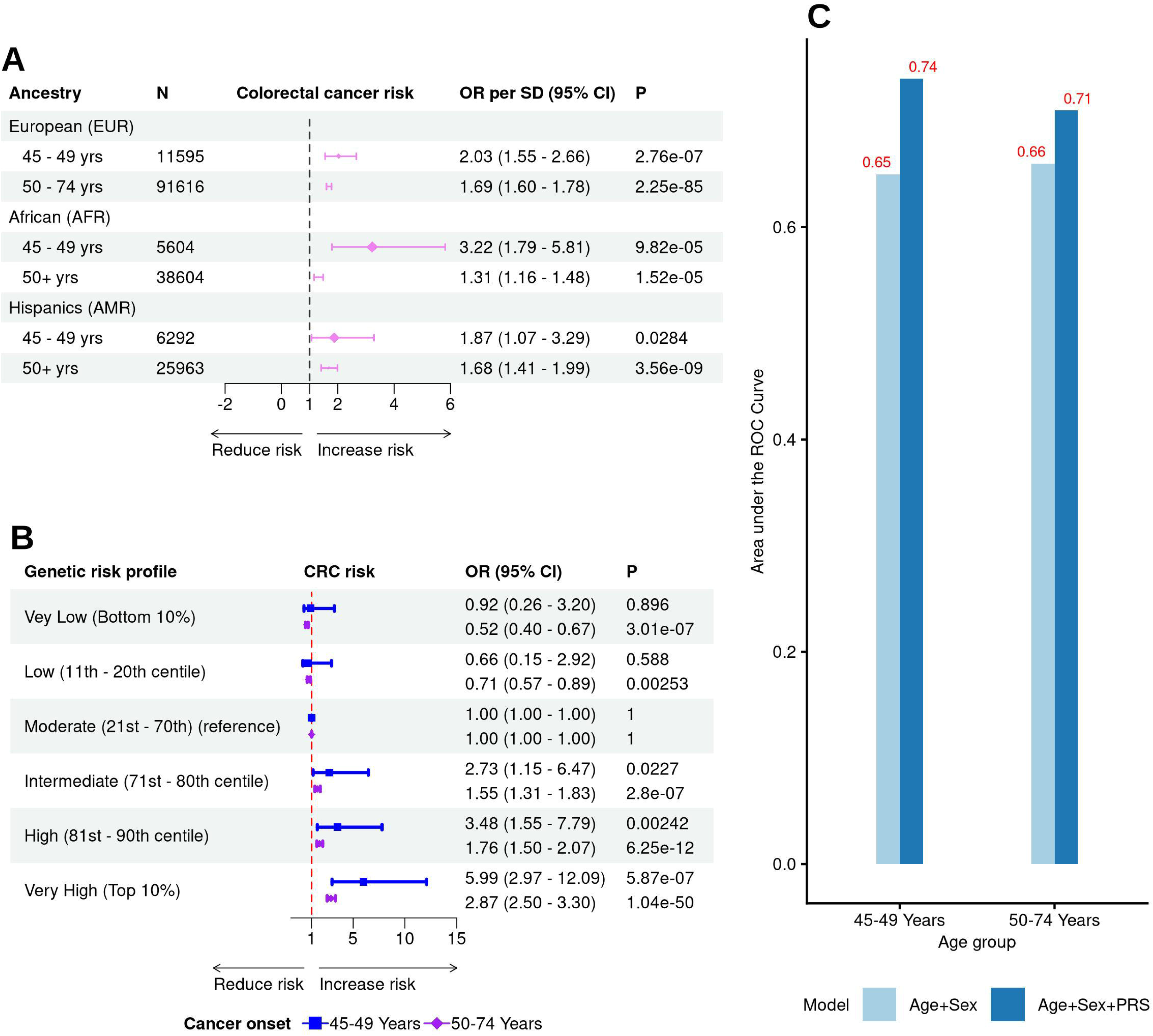
Comparison of the association and prediction of early and late onset of colorectal cancer. **Panel A**: The association of one standard deviation increase in the polygenic risk score (PRS) and the risk of early (45-49 years) and late (50+ years) onset of colorectal cancer in Europeans, African and Hispanics in the All of Us Cohort. The association is expressed in odds ratio (OR) per standard deviation increase in the genetic risk, and the 95% confidence interval. **B** - Comparison of the risk of CRC in 45-49 years old and 50-74 years old in Europeans in All of Us Cohort across different genetic risk strata (c.f. moderate risk group). **C** - Comparison of the predictive improvement based on the area under the receiver operating characteristic curve for both early (45-49 years) and late (50-74 years) onset of colorectal cancer in Europeans in the All of Us cohort.

The PRS improved prediction of EOCRC (45–49 years) by 9% beyond age, sex, and 10 PCs (AUC 0.74 vs 0.65, P = 0.012), exceeding the 5% improvement for LOCRC in EUR (AUC 0.71 vs 0.66, P < 2.2 x 10^−16^) (**Figure 6C; Supplementary Table 8**). In younger individuals (<50 years; N=62251; 117 cases), the PRS remained strongly associated with EOCRC (OR per SD = 1.92, 95% CI = 1.60–2.30, P = 3.40×10^−12^) (**Supplementary Table 10**) and improved prediction by 5% (AUC 0.79 vs 0.75, P=2.1×10^−3^) (**Supplementary Table 8**).

In AoU AFR (N=5604; 16 cases), a one–SD increase in PRS was associated with a 3-fold higher EOCRC risk at ages 45–49 (OR = 3.22, 95% CI = 1.79–5.81, P = 9.83×10□□), exceeding the association in AFR aged ≥50 (OR = 1.31, 95% CI = 1.16–1.48, P = 1.52×10□□) (**Figure 6A**). A similar pattern was observed in AMR (EOCRC OR = 1.87 vs LOCRC OR = 1.68).

## DISCUSSION

Using genetic and clinical data from 1.9 million individuals, we developed a robust CRC PRS with new construction methods. We demonstrated in six major ancestries its utility as a genetic screening tool to: identify high risk individuals with no family who would be otherwise ineligible for screening, reclassify individuals with family history for prioritized diagnostic screening, and identify younger individuals at the risk of EOCRC.

Genetic studies have largely focused on European populations, limiting PRS transferability to non-Europeans, where performance—especially in Africans—has been suboptimal[11]. We evaluated our PRS across AFR, AMR, EAS, SAS, and MID and found good performance overall, though modest in AFR, consistent with the need for ∼4-fold larger African GWAS to match European accuracy [32]. Notably, the PRS showed a strong association with EOCRC in AFR, with a threefold risk increase per SD. Overall, these results advance the universal applicability of genetic screening across ancestries.

Current CRC screening guidelines rely mainly on age and family history [3–6,30]. Adding the PRS significantly improved risk prediction across ancestries (except in AFR and MID, limited by case numbers), increasing AUC by up to 7%, 4%, 4%, and 5% in EUR, AMR, EAS, and SAS respectively—exceeding prior PRSs [8,9]. The PRS also improved prediction beyond age, sex, family history, smoking, and IBD by up to 6%, 3%, 6%, and 5% in EUR, AMR, EAS and SAS, respectively. These results highlight the value of polygenic screening done at any time in life for early CRC identification and personalized prevention.

We showed that even among individuals with affected FDRs, those in the bottom 20% PRS had risks comparable to or lower than average-risk individuals without family history and could be deprioritized for screening. Conversely, 20% without family history (top 20% PRS) had risks equal to or exceeding those with at least one affected FDR, and the top 30% and 10% mapped to Australian CRC screening guideline risk categories 2 and 3 [31]. These findings support initiating earlier screening and preventive interventions in genetically high-risk individuals. As high-risk individuals develop CRC 12–15 years earlier than average, screening could begin 12– 15 years earlier for those in the highest PRS centiles.

EOCRC incidence and mortality are rising [2], and polygenic screening could enhance prevention. National screening guidelines in Australia and USA allow individuals aged 45-49 to “opt in” for CRC screening [3–6]. In individuals aged 45–49, the PRS improved prediction twice as much as in those 50–74 (9% vs 5%) and showed stronger associations than for late-onset CRC across ancestries. Genetically high-risk individuals (top 10%) had a 6-fold higher EOCRC risk, comparable to those with affected FDRs and equivalent to high-risk categories (3) in the Australian CRC screening guidelines based on family history [31]. These results support prioritizing <50-year-olds with high PRS for earlier and more intensive screening and chemoprevention (e.g. low dose aspirin use).

Therefore, if genetic screening was to be incorporated in the current CRC screening guidelines e.g. for Australia, our study suggests that; i) iFOBT screening could be considered every two years from age 40 to 74 for those with high genetic risk but no family history. ii) Daily low dose aspirin could be considered in genetically high (>30%) individuals from age 40 (or 45years) to age 74 to reduce their risk of CRC. iii) Colonoscopy could be offered to individuals with high genetic risk every five years starting at age 50 to age 74. iv) Individuals with FDR FH, but low genetic risk could be screened less often or at the same rate as those with no FH but an average genetic risk.

This study has several strengths: a large sample enabling robust PRS development, use of advanced methods (SBayesRC and PRS-CSx) that incorporate millions of SNPs with important genetic information and cross-ancestry modeling; assessment across six ancestries (EUR, AMR, AFR, EAS, SAS, MID); and comprehensive analyses of clinically relevant scenarios, including early-onset CRC and family history. Study limitations include heterogeneous CRC definitions across cohorts, which may explain attenuated performance in AoU relative to UKB. Despite this, the PRS performed robustly, particularly. We could not exclude individuals with high-penetrance syndromes e.g. Lynch syndrome and familial adenomatous polyposis[33], though these account for only ∼5% of CRC [33], and were rare in AoU. That is, only 40 (0.01%) and 20 (0.006%) participants out of 354,400 were reported to have Lynch syndrome and familial adenomatous polyposis in AoU, respectively. Thus, limiting their impact in our study.

## Conclusion

Our findings provide comprehensive evidence to support potential integration of genetic screening into clinical practice guidelines for CRC screening, early identification and prevention.

## Data availability

The CRC PRS weights will be publicly available in the Polygenic Score Catalog upon publication. Researchers interested in accessing the following biobanks can apply via the respective study websites; UKB (https://www.ukbiobank.ac.uk/enable-your-research/apply-for-access), CARTaGENE (https://cartagene.qc.ca), and CLSA (https://www.clsa-elcv.ca/). Researchers interested in collaborating with QSkin should contact the principal investigator at. The GWAS summary on CRC from Fernandez-Rozadilla (GWAS Catalog GCST90255676), FinnGen (https://www.finngen.fi/en/access_results), Million Veteran Program (via dbGaP at https://ftp.ncbi.nlm.nih.gov/dbgap/studies/, under accession number accession number phs002453), the EAS GWAS meta-analysis data between BBJ and NBK were accessed via https://koges.leelabsg.org/pheno/META_COLCA. This study used data from the All of Us Research Program’s Controlled Tier Dataset version 8, available to authorized users on the Researcher Workbench (https://allofus.nih.gov/article/opportunities-researchers).

## Acknowledgments

The authors would like to thank the study participants in the All of Us, UKB, CARTaGENE, QSkin, CLSA, AoU, MVP, FinnGen and other previous studies in the GECCO and CORRECT Consortia and Japan Biobank and Korean Biobank who made this study possible. This study was conducted using data from the UK Biobank (application number 25331). We also thank the National Institutes of Health’s All of Us Research Program for making the participant level data available for this study. This research has been conducted using de-identified data from the CARTaGENE Biobank (project ID: 488635). CARTaGENE obtained ethics approval from the CHU Sainte-Justine (reference number MP-21-2011-345, 3297). Written consent was acquired from all participants in CARTaGENE. We thank the participants and investigators of the CARTaGENE study.

This research was made possible using the data/biospecimens collected by the Canadian Longitudinal Study on Aging (CLSA). Funding for the Canadian Longitudinal Study on Aging (CLSA) is provided by the Government of Canada through the Canadian Institutes of Health Research (CIHR) under grant reference: LSA 94473 and the Canada Foundation for Innovation, as well as the following provinces, Newfoundland, Nova Scotia, Quebec, Ontario, Manitoba, Alberta, and British Columbia. This research has been conducted using the CLSA dataset [Baseline Comprehensive Dataset version 4.0, Follow-up 1 Comprehensive Dataset version 1.0, March 2021 (Version 3) release of genetic data], under Application Number 190225. The CLSA is led by Drs. Parminder Raina, Christina Wolfson and Susan Kirkland. The time and commitment of the participants to the CLSA study platform is gratefully acknowledged, without whom this research would not be possible.

## Patient consent for publication

Not applicable

## Ethics approval

The human research ethics committee at QIMR Berghofer Medical Research Institute provided ethical oversight to this study (P3501). All participants in the contributing cohorts provided informed consent to participate in this study.

## CrediT Authorship Contributions

Abishna Parasuraman, BSc(Hon) (Formal analysis: Equal; Investigation: Equal; Project administration: Equal; Visualisation: Equal; Writing – original draft: Supporting; Writing – review & editing: Equal)

Amanda Wei-Yin Lim, BPharm, MSc (Formal analysis: Supporting; Investigation: Supporting; Project administration: Supporting; Writing – review & editing: Equal)

Rawaa Eltayib, MD, MSc (Formal analysis: Supporting; Investigation: Supporting; Project administration: Equal; Writing – review & editing: Supporting)

Nirmala Pandeya, PhD (Data curation: Equal; Resources: Equal; Writing – review & editing: Equal)

Catherine M Olsen, PhD (Data curation: Equal; Resources: Equal; Writing – review & editing: Equal)

Graham Radford-Smith, MD, PhD (Writing – review & editing: Equal)

David C. Whiteman, MBBS(Hons), PhD (Funding acquisition: Equal; Resources: Equal; Writing – review & editing: Equal)

Stuart MacGregor, PhD (Conceptualization: Lead; Funding acquisition: Lead; Investigation: Lead; Methodology: Lead; Resources: Lead; Supervision: Lead; Writing – review & editing: Lead)

Mathias Seviiri, MD, MPH, PhD (Conceptualization: Lead; Data curation: Lead; Formal analysis: Lead; Funding acquisition: Lead; Investigation: Lead; Methodology: Lead; Project administration: Lead; Resources: Lead; Visualisation: Lead; Software: Lead; Supervision: Lead; Writing – original draft: Lead; Writing – review & editing: Lead)

