## Supplementary Methods for "Genetic prediction of colorectal cancer risk in six major ancestries provides insights to streamline practice screening guidelines"

**Author affiliations list**

1. Department of Population Health, QIMR Berghofer, Brisbane, Australia
2. Faculty of Health, Medicine and Behavioural Sciences, The University of Queensland, Brisbane, Queensland, Australia
3. Department of Gastroenterology and Hepatology, Royal Brisbane and Women's Hospital, Brisbane, Queensland, Australia
4. Gut Health, QIMR Berghofer, Brisbane, Queensland, Australia.
5. School of Biomedical Sciences, Faculty of Health, Queensland University of Technology, Brisbane, Queensland, Australia

* Corresponding author:
Mathias Seviiri, Statistical Genetics, Department of Population Health, QIMR Berghofer, Brisbane, Australia.
300 Herston Road, Herston, Brisbane, Queensland 4006, Australia

### Supplementary Methods

##### Cohorts for evaluating the performance of the polygenic risk score

#### The UK Biobank, the United Kingdom

The UK Biobank is a large longitudinal population-based study in the United Kingdom that recruited ~500,000 participants aged 40-69 years at the time between 2006 and 2010 [1,2]. Further details on the recruitment process, data processing and phenotyping have been previously published [1,2]. In the UK Biobank we selected a holdout dataset with 5 major ancestries including Europeans (EUR), Africans (AFR; N=7029), Hispanics (AMR; N=2,465), East Asians (EAS; N=1,930) and South Asians (SAS; N=7460). The EUR dataset comprised 10% of EUR participants in UKB (N=34079), randomly selected and not related to each other or to the 90% who were in the discovery cohort. All participants were at least 75% genetically similar to their respective ancestry in 1000 Genome Phase 3 super populations [3]. CRC cases were based on ICD 10 and 9 codes, while controls had no history of any cancer. Cases were ascertained through data linkage with the Cancer Registry in the UK. Data were also curated on first degree relative family history of CRC, colorectal polyps, colorectal benign neoplasms, inflammatory bowel disease (IBD), and smoking status. Baseline characteristics of the participants for each ancestry are summarised in **Supplementary Table 2).** The National North West Multi-Centre Research Ethics Committee in the United Kingdom approved the study protocol (ref. 11/NW/0382). All participants provided written informed consent to participate in the study.

##### The QSkin Sun and Health Study (QSkin) Cohort, Australia

The QSkin Sun and Health Study (QSkin) is a longitudinal population-based cohort (N~43,000) of adult participants recruited between 20101and 2011 in Queensland, Australia. Details on participant recruitment, data processing and quality control processes have been published [4]. In brief, at the time of recruitment, participants were aged 40-69 years. Data were collected in 2 phases; QSkin 1 in 2011 (N ~ 17,000 with both genetic and clinical data), then followed by QSkin 2 in 2019 (N ~ 8,000 with genetic and clinical data). Participants were genotyped using the Illumina GSA SNP chip. Standard genetic quality control [5] was performed on both samples and single nucleotide polymorphism (SNPs). Genetic information was imputed to the TOPMed reference panel [6]. CRC cases were ascertained through data linkage to the Australian Cancer Database. Controls were participants with no history of CRC. The human research ethics committee at QIMR provided ethical oversight for the study. For these analyses, the study sample was restricted to 23,129 (336 cases) participants who provided written informed consent to be included in study and had genetic data. Baseline characteristics of the participants are summarised in **Supplementary Table 2.**

**The All of Us Research Program, the United States of America**

The All of Us Research Program (AoU) is a longitudinal cohort study in the United States of America with a target to enroll about 1 million participants of diverse backgrounds. Further information on participant recruitment, data collection, processing, quality control measures, data types, and linkages have been published before [7,8]. Participant information at baseline is summarised in **Supplementary Table 2.** In order to test the PRS across multiple ancestries, we also applied the PRS in the AoU cohort after conducting a leave-one out meta-analysis discovery GWAS that excluded AoU participants. We applied PRS in the AoU V8 (after new data release by AoU) participants who were not related, passed quality control checks, not sex discordant, and had short read whole genome sequence data. Participants were EUR (N=153,867), AFR (N=65621), AMR (N=65,252), EAS (N=8495) and Middle Easterns (MID; N=1231). Baseline characteristics of the participants are summarised in **Supplementary Table 2.** We also selected a subpopulation of participants aged 45 - 49 years to allow analysis for early onset of CRC; EUR (N=11,595), AFR (N=5,604) and AMR (N=6,292). Cases were based on ICD 9 (153 and 154) and ICD10 (C18, C19, and C20) codes, with at least 2 EHRs, while controls were participants with no history of CRC. We also curated data on first degree family history of CRC, smoking, and IBD diagnosis. The AoU Institutional Review Board (IRB) provided ethical oversight over the study, and all participants provided informed consent.

**CARTaGENE**, **Canada**

The **CARTaGENE** study is a large, population-based and prospective cohort of adult participants in Quebec, Canada. Genetic, clinical/health and lifestyle data were collected from 43,000 participants aged between 40 and 69 years at recruitment. Detailed information on participant recruitment, data collection and processing, genotyping and genotyping QC have been published [9,10]. In brief, participants were recruited in two phases; Phase A between 2009 and 2010 for 19,784 participants, and Phase B between 2012 and 2015 for an additional 23,000 participants [9,10]. At baseline, participants had their information recorded including age, sex, presence of chronic conditions (e.g cancer of colon or rectum), and familial history of neoplasms (e.g. cancer of colon or rectum) [9]. CRC cases were ascertained through a self reported survey. Controls were participants with no history of any cancer. Participants were genotyped using the Illumina GSA SNP chip, and after genotyping QC, they were imputed to the TOPMed panel [6]. In the present analysis, we computed the PCs in CARTaGENE, and projected them to the 1000 Genomes Phase 3 [3]. We then used the random forest classifier to ascertain the predicted genetic ancestry of all the samples. We restricted our analysis to 23,320 participants who were at least 60% genetically similar to the EUR in 1000 Genomes. Baseline characteristics of the participants are summarised in **Supplementary Table 2.** The research ethics board of the Sainte-Justine University Health Center provided ethical oversight to CARTaGENE, and participants provided informed consent to participate in the study.

**Performing and collating the discovery GWAS for the SBayesRC PRS models**

We performed a large-scale trans-biobank meta-analysis GWAS on CRC in EUR (N=1,448,354; 103,401 cases) (**Figure 1 and Supplementary Table 1)**. In the AoU biobank, we conducted case-control GWAS on CRC in EUR (N=12,838; 1,617 cases), using short-read whole genome sequence data in the Curated Data Repository release 7 (CDR V7). CRC cases were based on EHR. Controls were individuals with no history of CRC. We also conducted case-control GWAS on colorectal polyps, and colorectal benign neoplasms in the EUR dataset, to enable us to subsequently conduct multi-trait GWAS (**Supplementary Table 1).** Regenie V3.2.6 [11] was used to conduct all analyses, and we adjusted for age, sex, and 10 PCs for each GWAS.

In the UK Biobank, first we conducted case-control GWAS for CRC in the EUR dataset (N=341316; 8523 cases) using phenotype data from February 2024. Age, sex, and 10 PCs were adjusted for in the analysis. In the UKB, CRC cases were determined through data linkage to cancer registries. We defined CRC cases based on ICD10 codes (data field 40006); C18, C19, and C20 and ICD9 codes (data field 40013); 153 and 154. Controls were participants with no history of any cancer. In addition, we conducted case-control GWASs on colorectal polyps (N=448613; 36280 cases) and colorectal benign neoplasms (N=454289; 30881 cases) in EUR. colorectal polyp cases were based on ICD10 codes; K635 and K621, while benign neoplasms of colon and rectum cases were defined by ICD10 codes D12 (data field 41270). Controls for colorectal polyps and colorectal benign neoplasms excluded the respective cases as well as participants with CRC. We used SAIGE [12] to conduct all the GWAS and we controlled for age, sex, and 10 PCs.

Second, for purposes of validating the PRS in EUR in the UK Biobank, we split the UKB EUR dataset into 2 independent datasets. We performed case-control GWAS for CRC in the selected 90% EUR dataset (N=307146; cases=7671, and controls = 299475). We reserved an independent dataset (10% EUR) for validation of the PRS. In a similar way, we conducted CRC GWASs for colorectal polyps (N=415047; 33695 cases) and colorectal benign neoplasms (N=414844; 28724 cases) in the 90% EUR dataset.

In CLSA, we performed a CRC case-control GWAS (N=17442; cases = 351). Cases were self-reported CRC diagnoses, while controls were participants with no history of CRC. SAIGE [12] was used to conduct the analysis, and we controlled for age, sex, and 10 PCs.

We also accessed GWAS summary statistics on CRC (N=353919; 8801 cases) from FinnGen [13], based on data freeze 11. FinnGen is a large biobank with genetic and clinical data on over 500,000 Finnish residents [13]. Cases were based on ICD10 codes, while controls had no history of any cancer diagnosis. The FinnGen analysis was conducted using SAIGE [12]. GWAS summary statistics on colorectal adenoma (N=349265; 4147 cases) and colorectal benign neoplasms (N=453733; 25190 cases) were also accessed from FinnGen.

In addition, we collated recent GWAS summary data from the MVP on CRC in EUR (N=446766; 10436 cases), AFR (N=120570; 2466 cases) and AMR (N=59419; 1003 cases) [14]. MVP is a large-scale prospective cohort study that is investigating how genes impact diseases e.g. cancers among veterans in the USA. We accessed GWAS data based on release 5 [14], conducted using SAIGE [12]. In MVP CRC cases were determined by ICD codes from the EHR. We also accessed GWAS data on colorectal polyps (N=315668; 73737 cases), and colorectal benign neoplasms (N=395250; 125095 cases) for the EUR descent participants. We accessed the GWAS summary statistics for recently published findings on CRC in EUR that excluded UKB (GCST90255676; N=160,527; 73,673 cases) [15].

After collating GWAS summary statistics from multiple biobanks and consortia (as described above), therefore we performed a large meta-analysis GWAS on CRC in EUR (N=1,448,354; 103,401 cases) using GWAS summary data from AoU, UKB, MVP, FinnGen, CLSA and Fernandez-Rozadilla et al 2023 (GECCO and CORECT consortia) (**Supplementary Table 1**). We applied the inverse-variance-weighted fixed-effects model implemented in metal [16]. We retained SNPs with a minor allele frequency of at least 1% and present in at least 2 contributing cohorts. The GWAS summary statistics were used as input for the **SBayesRC PRS models (**Refer to **Materials and Methods: Statistical analysis - PRS construction approach 1: SBayesRC)**

**Performing and collating the discovery GWAS for the PRS-CSx PRS models**

We used LD score regression [17] to quantify the genetic correlation between CRC, colorectal polyps, and colorectal benign neoplasms in EUR. Next, we performed a multi-trait GWAS using MTAG [18] incorporating CRC (N=1,448,354; 103,401 cases), colorectal polyps (N=1,241,930; 125,589 cases), and colorectal benign neoplasms (N=1,431,656; 198,837 cases) in EUR participants. (**Supplementary Table 1**). The colorectal polyps input was a large meta-analysis GWAS for EUR data from MVP, AoU, and UKB, whilst the colorectal benign neoplasms included data from FinnGen, MVP, AoU, and UKB.

In order to construct a trans-ancestry PRS including information from participants of East Asian descent, we accessed publicly available GWAS meta-analysis data of CRC (N=239989; 8545 cases) based on analysis in BBJ, and NBK[19]. Next, using the data from the BBJ-NBK meta-analysis, and GWAS summary data from CKB (N=75947; 434 cases) we performed an IVW fixed effects meta-analysis GWAS of EAS (N=315,936; 8979 cases) using metal [16]. Next, we collated recently released GWAS summary statistics on CRC for AFR (N=120,570; 2,466 cases), and AMR (N=59,419; 1,003 cases) from MVP [14].

Lastly, we used the European MTAG [18] summary statistics for CRC together with the GWAS data for AFR, AMR, and EAS to construct a trans-ancestry PRS using PRS-CSx [20], which jointly models ancestry-specific LD and effects sizes to improve cross-population prediction. Based on PRS weights from PRS-CSx, PLINK 1.9/2.000 [21] was used to generate the raw PRS scores for each individual in the validation datasets of non-European ancestry in the UKB, and AoU.
